# Preload-Afterload Failure Patterns During Human Defecation: Mechanistic Phenotyping and Reference Limits

**DOI:** 10.64898/2026.11.27.26369117

**Authors:** Daming Sun, Jifan Yang, Lei Ke, Hans Gregersen

## Abstract

**Background:** Rear and front Fecobionics pressures describe propulsive preload and outlet afterload throughout evacuation. We tested whether complete preload-afterload trajectories reveal distinct failure mechanisms in fecal incontinence (FI) and defecatory disorders (DD).

**Methods:** We retrospectively analyzed 93 examinations with usable paired pressures: 22 healthy participants (HP), 38 FI, and 33 DD. Pressure-space work rate, the primary endpoint, was the absolute front-pressure line integral with respect to rear pressure divided by evacuation duration. Secondary measures described pressure separation, preload, release, cycling, and developmental healthy reference limits.

**Results:** The median pressure-space work rate was 803.5 cmH_2_O^2^/s in HP, 389.3 in FI, and 528.8 in DD (P=0.0061). FI differed from HP after Holm correction (P=0.0090; area under the curve 0.732). FI also showed lower peak rear-front gradient, rear preload, and front-pressure release. DD group averages overlapped HP, but lower work rate, rear pressure, and gradient correlated with worse constipation scores (rho=-0.63 to -0.66; adjusted P<0.001). DD patients unable to complete anorectal-manometry expulsion had lower gradients than timed completers (69.9 versus 155.4 cmH_2_O; adjusted P=0.025). Within FI, work output correlated with resting but not squeeze pressure, leaving both continence-system impairment and anticipatory suppression of abdominal effort as plausible mechanisms.

**Conclusions:** FI was characterized by reduced pressure-work output, whereas DD contained heterogeneous preload, afterload, release, and repetitive-loading patterns. Trajectory-based phenotyping warrants prospective validation.

**New & Noteworthy:** Preload-afterload trajectories may reveal failure mechanisms in fecal incontinence (FI) and defecatory disorders (DD). DD showed heterogeneous preload, afterload, release, and repetitive-loading patterns. Lower work rate, rear pressure, and gradient correlated with worse constipation scores. Non-evacuators had lower gradients than evacuators. FI showed lower pressure-space work rate, peak rear-front gradient, preload, and front-pressure release. FI work output correlated with resting but not squeeze pressure, leaving continence-system impairment and anticipatory abdominal effort suppression as plausible mechanisms.

## Introduction

Defecation requires the coordinated generation of propulsive pressure and reduction of outlet loading as rectal contents traverse the anorectum. Its success therefore depends on the interaction between propulsion and outlet opening throughout an evacuation attempt. Simultaneous measurements of anorectal pressures, abdominal and anorectal motion, and evacuation have demonstrated that normal defecation involves a coordinated sequence of pressurization and subsequent anal relaxation, whereas impaired evacuation can involve inadequate pressurization, insufficient relaxation, or disturbed coordination. ^1–10^ A physiological assessment should consequently characterize both the magnitude of loading and its relationship to outlet unloading over time.

Established manometric classifications for defecatory disorder (DD), including the Rao dyssynergia patterns, recognize inadequate propulsion, impaired anal relaxation, and paradoxical contraction. ^4–5^ However, a pressure pattern observed during simulated defecation does not necessarily identify the process limiting evacuation: patterns conventionally classified as dyssynergic also occur in asymptomatic individuals. This creates a need to examine how pressure generation and outlet loading evolve during passage of a stool-like object and whether their interaction relates to symptoms and evacuation performance.

Fecobionics records pressures at the rear and front of a simulated stool during attempted evacuation. Plotting front pressure against rear pressure represents the evolving relationship between loading behind the device and loading ahead of it. We use preload and afterload as operational descriptions of these pressure signals rather than direct measurements of muscle length, propulsive force, or outlet resistance. ^11–15^ The trajectories permit examination of whether front pressure decreases while rear pressure remains elevated, whether an adverse pressure relationship persists during effort, and whether repeated loading occurs without sustained unloading. ^11–15^ Although earlier Fecobionics studies have described disease-associated pressure patterns and dyssynergia phenotypes, ^11–27^ the relationship of quantitative trajectory measures to symptom severity and independent evacuation performance remains incompletely defined.

Fecal incontinence (FI) presents a complementary physiological question. Although principally a disorder of continence,^1,4,8,9^ impaired outlet closure could influence the pressure strategy adopted during intentional evacuation. Reduced pressure output might reflect altered sensorimotor function or pressure transmission, reduced recruitment of propulsive effort, or a learned strategy that limits abdominal pressurization when increased pressure is associated with leakage. These explanations predict similar habitual pressure patterns but different responses to standardized coaching and maximal-effort testing.

We therefore examined whether complete rear–front pressure trajectories identify clinically relevant differences in propulsion–outlet interaction in healthy participants (HP) and patients with FI or DD. We hypothesized that FI would exhibit reduced pressure-trajectory output, whereas DD would show heterogeneous disturbances in loading and unloading, with less favorable pressure relationships accompanying greater constipation severity and impaired evacuation. In additional exploratory analyses, we assessed whether pressure-space work rate contributed information about constipation severity beyond peak rear pressure and evacuation duration. Developmental reference limits were used to describe individual departures from healthy behavior. The study was intended to identify candidate physiological mechanisms and testable predictions rather than establish their neural, muscular, or behavioral causes.

## Methods

### Study design and participants

This retrospective secondary analysis used baseline Fecobionics examinations performed at the Department of Surgery, the Chinese University of Hong Kong. Participants were classified from the original clinical records as healthy participants (HP), FI, or DD/constipation using the Rome Criteria, symptom scores and relevant testing of function and anatomy such as anorectal manometry (ARM), balloon expulsion test (BET), defecography and endoanal ultrasonography.^12,14,15,28^ Patients appearing in more than one source workbook were linked by study identifier and counted only once. Patients with mixed FI-DD group were not considered in the data material. The study was IRB approved (protocol no. 2017.122, Joint CUHK-NT East Cluster Clinical Research Ethics Committee). All experiments were performed in accordance with relevant guidelines and regulations. Trial Registration. https://www.clinicaltrials.gov ID: NCT03317938. Date of registration: 23/10/2017.

The expanded cohort was prespecified as primary because it maximized objectively recoverable examinations without relying on manually inferred terminal intervals. The strict cohort retained conventionally marked intervals only. The maximum sensitivity cohort additionally included conservatively reviewed terminal passage intervals. No important hardware, software, or protocol changes occurred during recruitment.

### Signal processing and interval selection

Raw files contained time, front pressure, bag pressure, rear pressure, bend angle, and front and rear orientation. Pressures were corrected by subtracting the median during the 20 seconds before the analyzed interval. Split or restarted recordings were retained when the selected interval and its baseline were intact. Sustained pressure elevations and long repetitive contractions were preserved. Isolated extreme front-channel spikes lasting no more than 0.25 seconds were interpolated only when adjacent samples returned to the physiological range. Short-window median smoothing was used only for trajectory landmarks and contraction counting. A failed front channel excluded that examination from paired-pressure analysis but not from studies of its other valid channels.

### Preload-afterload variables

Rear pressure was treated as preload and front pressure as afterload. The line front pressure=rear pressure represented pressure unity; front pressure at or above rear pressure during active effort represented an adverse instantaneous gradient. Pressure-space work was defined as a pressure-plane line integral and has units of pressure squared. It is a trajectory descriptor analogous to cyclic work, not physical work in joules. Full equations are given in Supplementary Methods.

The primary endpoint was pressure-space work rate: the absolute line integral of front pressure with respect to rear pressure over the evacuation trajectory divided by interval duration. Key secondary endpoints were absolute pressure-space work, peak rear-front gradient, peak rear pressure, and front-pressure release amplitude (see Supplementary Methods). Exploratory variables included signed work, hysteresis ratio, phase-path length, active-afterload fraction, loading and release slopes, pressure cycling, contraction count, and front-peak timing.

### Developmental reference phenotypes

The normative subset excluded individuals who were only conditionally labeled as healthy, along with healthy participants who reported unusual symptom scores. Fifth and 95th percentiles defined high-specificity developmental limits. Inadequate preload was rear peak below the fifth percentile. Excessive afterload was active-effort time with front pressure at or above rear pressure above the 95th percentile. Impaired release was front-pressure release below the fifth percentile or terminal front residual above the 95th percentile. Repetitive loading was pressure cycling or contraction count above the 95th percentile. Categories could overlap; more than one abnormal mechanism constituted mixed failure.

### Clinical variables and competing FI mechanisms

ARM-BET resting pressure, maximum squeeze pressure, and expulsion time were obtained. A blank expulsion time accompanied by “cannot evacuate within 120 seconds” was coded as categorical non-expulsion rather than missing at random. Associations of FI pressure output with resting and squeeze pressure were examined to distinguish, incompletely, continence-system impairment from generalized muscle capacity. Behavioral avoidance, fear of leakage, and other psychological factors were not directly measured; consequently, anticipatory suppression remained an explanatory hypothesis rather than an observed exposure.^8–10,29^

### Statistical analysis

Continuous variables are reported as median (interquartile range). Three groups were compared with Kruskal-Wallis tests. Prespecified pairwise Mann-Whitney tests used Holm adjustment within each endpoint. Directionless receiver operating characteristic areas quantified discrimination. Spearman correlations tested construct validity; Holm adjustment was applied within each outcome family. Bootstrap intervals used 2,000 or 3,000 resamples. Robustness was evaluated by first/99th-percentile winsorization, five-percent winsorization for correlations, leave-one-out analyses, and repetition in strict and maximum cohorts. Reference-percentile uncertainty used 4,000 bootstrap resamples. Statistical outliers were not deleted.

Exploratory incremental analyses in DD assessed the association between pressure-space work rate and constipation severity after adjustment for peak rear pressure and evacuation duration using partial Spearman correlation, participant-bootstrap confidence intervals, and residual-permutation testing. Leave-one-out cross-validation compared prediction of constipation scores from linear models containing log-transformed rear pressure and duration with otherwise identical models additionally containing log-transformed work rate. Sensitivity analyses incorporated front-pressure amplitude or substituted peak gradient for rear pressure and repeated the comparisons across cohort definitions.

## Results

### Cohort and data quality

Paired front-rear pressure analysis included 93 examinations: 22 HP, 38 with FI, and 33 with DD **(Table 1).** The strict cohort contained 85 and the maximum cohort 99 examinations. One DD examination available for non-front-channel analyses was excluded because the front channel failed. The normative reference subset contained 20 HP. Fifteen boundary or extreme trajectories underwent diagnosis-blinded technical review. Three contained isolated spikes in bag, rear, or front pressure, but none justified subject exclusion; physiological adjudication remains pending. **Figure 1** shows representative examples of preload-afterload data from HP, FI and DD.

**Table 1.** Cohort characteristics. Values are median (interquartile range) unless otherwise indicated.

| **Characteristic** | **Healthy participants** | **Fecal incontinence** | **Defecatory defecation** |
|---|---|---|---|
| Participants, n | 22 | 38 | 33 |
| Age, years | 56.0 (47.2-65.2) | 61.5 (58.0-70.5) | 58.0 (42.0-67.0) |
| Women, n (%) | 15 (68%) | 33 (87%) | 26 (79%) |
| Conventional marked interval, n (%) | 19 (86%) | 38 (100%) | 28 (85%) |

**Figure 1.**
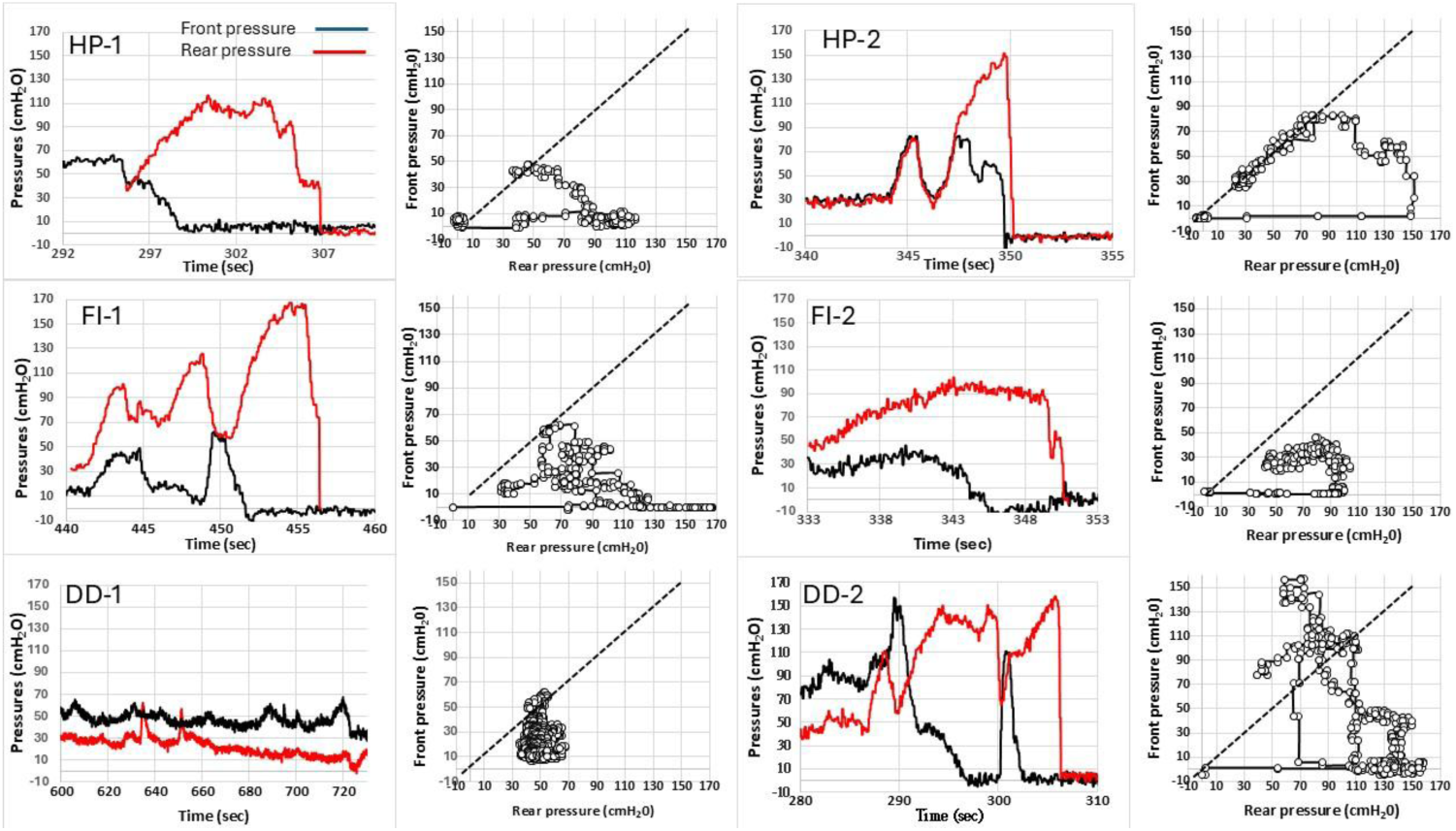
Representative pressure-time tracings and preload-afterload trajectories. The dashed pressure-unity line marks equal front and rear pressures; points above it have an adverse instantaneous gradient. Two examples are shown from each group, e.g., for the defecatory disorder (DD) patients, one example is a low-pressure non-evacuator whereas the other clearly show dyssynergic pattern with paradoxical contractions and reverse loops. Healthy participants and fecal incontinence are abbreviated HP and FI.

### Primary pressure-space work analysis

Median pressure-space work rate was 803.5 cmH2O^2^/s in HP, 389.3 in FI, and 528.8 in DD (overall P=0.0061; **Table 2 and Figure 2**). FI was lower than HP evacuation (Holm-adjusted P=0.0090) and DD (P=0.0368), whereas DD did not differ from HP (P=0.5766). FI-versus-HP discrimination was 0.732 (bootstrap interval 0.591-0.862). Absolute pressure-space work showed the same low-output FI pattern and was higher in DD than FI.

**Table 2.** Primary and key secondary preload-afterload measures. Values are median (interquartile range).

| **Measure** | **Healthy participants (HP)** | **Fecal incontinence (FI)** | **Defecatory defecation (DD)** | **Overall P** | **HP vs FI, adjusted P** | **HP vs DD, adjusted P** | **FI vs DD, adjusted P** |
|---|---|---|---|---|---|---|---|
| Pressure-space work rate, cmH<sub>2</sub>O<sup>2</sup>/s | 803.5 (523.3-1135.8) | 389.3 (185.0-623.0) | 528.8 (326.0-1360.7) | 0.0061 | 0.0090 | 0.5766 | 0.0368 |
| Absolute pressure-space work, cmH<sub>2</sub>O<sup>2</sup> | 19316.4 (12189.3-39591.8) | 11586.3 (4911.8-19894.3) | 23227.7 (13085.1-50229.8) | 0.0048 | 0.0265 | 0.9111 | 0.0097 |
| Peak rear-front gradient, cmH<sub>2</sub>O | 140.4 (119.4-186.4) | 95.8 (55.6-134.1) | 134.7 (79.5-177.1) | 0.0043 | 0.0027 | 0.3067 | 0.0873 |
| Peak rear pressure, cmH<sub>2</sub>O | 112.8 (93.9-146.3) | 67.5 (42.0-117.3) | 107.6 (47.5-143.8) | 0.0320 | 0.0177 | 0.4566 | 0.4566 |
| Front-pressure release amplitude, cmH<sub>2</sub>O | 98.4 (61.6-130.5) | 58.6 (33.7-88.4) | 85.4 (58.7-102.8) | 0.0066 | 0.0239 | 0.3360 | 0.0239 |

**Figure 2.**
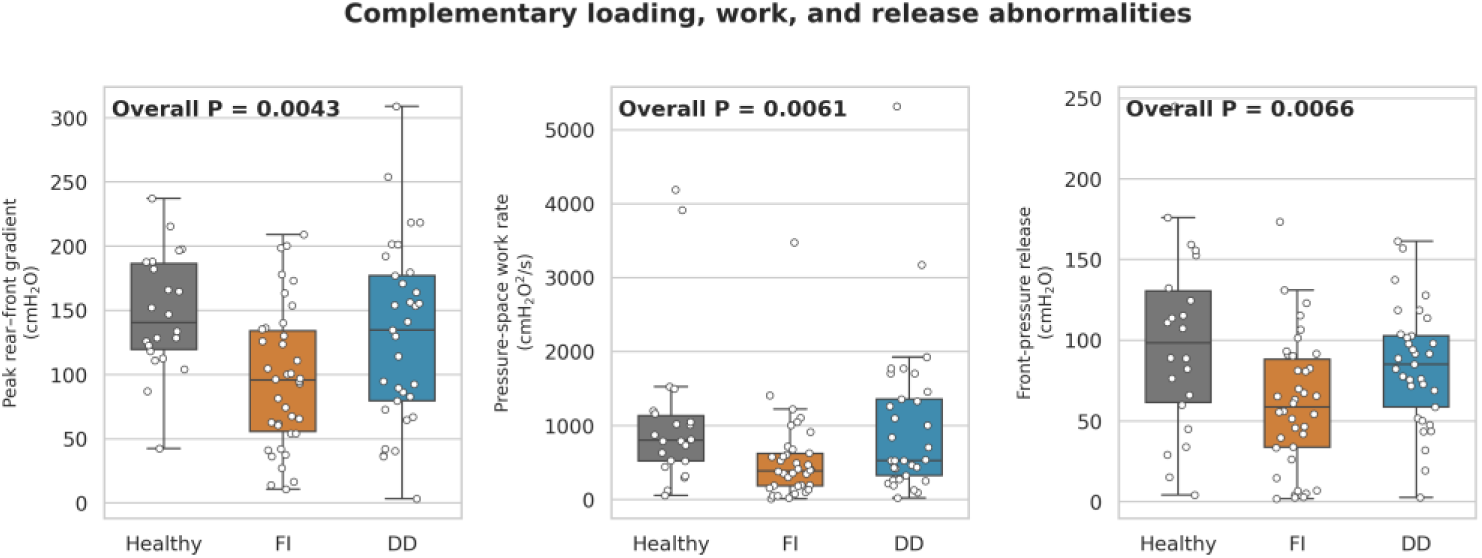
Complementary loading, pressure-space work, and release abnormalities. Boxes show medians and interquartile ranges; points show individual participants. P values are three-group Kruskal-Wallis tests. Defecatory disorder and fecal incontinence are abbreviated DD and FI.

The primary inference persisted across cohort definitions. Median work rate was 813.7, 389.3, and 622.9 cmH2O^2^/s in the strict HP, FI, and DD groups (P=0.0017), and 803.5, 399.7, and 526.9 in the maximum cohort (P=0.0139). Rank-test inference was unchanged by first/99th-percentile winsorization.

### Loading, gradient, and release

FI also showed reduced peak rear-front gradient, peak rear preload, and front-pressure release **(Table 2).** Median gradients were 140.4 cmH_2_O in HP, 95.8 in FI, and 134.7 in DD (P=0.0043). HP versus FI remained significant after Holm correction (P=0.0027; area under the curve 0.760). Front-pressure release was 98.4, 58.6, and 85.4 cmH_2_O, respectively (P=0.0066). Representative trajectories illustrate coupled loading followed by release in health, low-amplitude cycling in FI, and heterogeneous repeated or adverse-afterload paths in DD (**Figure 1).**

### Mechanistic reference patterns

Using developmental reference limits, inadequate preload occurred in 4.5% of HP, 28.9% of FI, and 21.2% of DD examinations. Excessive afterload occurred in 4.5%, 10.5%, and 21.2%; impaired release in 9.1%, 26.3%, and 18.2%; and repetitive loading in 9.1%, 7.9%, and 15.2% **(Table 3**; **Figures 3 and 4).** Overall prevalence differences were not statistically significant. These categories therefore describe trajectories outside developmental healthy reference limits rather than validated disease classifiers. Bootstrap intervals around the healthy limits were wide, particularly for cycling variables.

**Table 3.** Developmental mechanistic patterns. Categories may overlap; P values compare all three groups.

| **Mechanism** | **Healthy participants** | **Fecal incontinence** | **Defecatory defecation** | **Overall P** |
|---|---|---|---|---|
| Inadequate preload | 1/22 (4.5%) | 11/38 (28.9%) | 7/33 (21.2%) | 0.077 |
| Excessive afterload | 1/22 (4.5%) | 4/38 (10.5%) | 7/33 (21.2%) | 0.166 |
| Impaired release | 2/22 (9.1%) | 10/38 (26.3%) | 6/33 (18.2%) | 0.260 |
| Repetitive loading | 2/22 (9.1%) | 3/38 (7.9%) | 5/33 (15.2%) | 0.591 |

**Figure 3.**
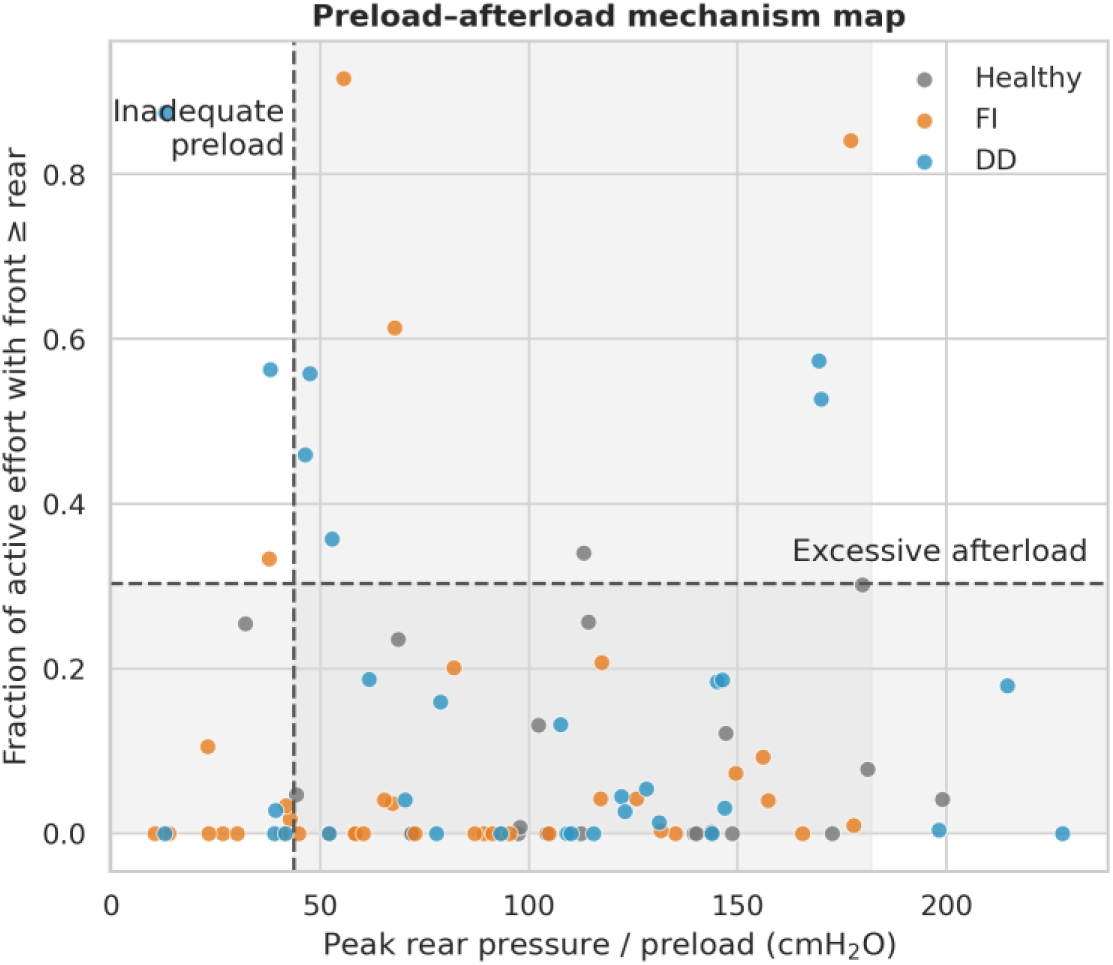
Preload-afterload mechanism map. Dashed lines show developmental fifth-percentile preload and 95th-percentile active-afterload limits from the normative healthy subset. Shading indicates the univariate reference bands; limits are not validated diagnostic cutoffs. Defecatory disorder and fecal incontinence are abbreviated DD and FI.

**Figure 4.**
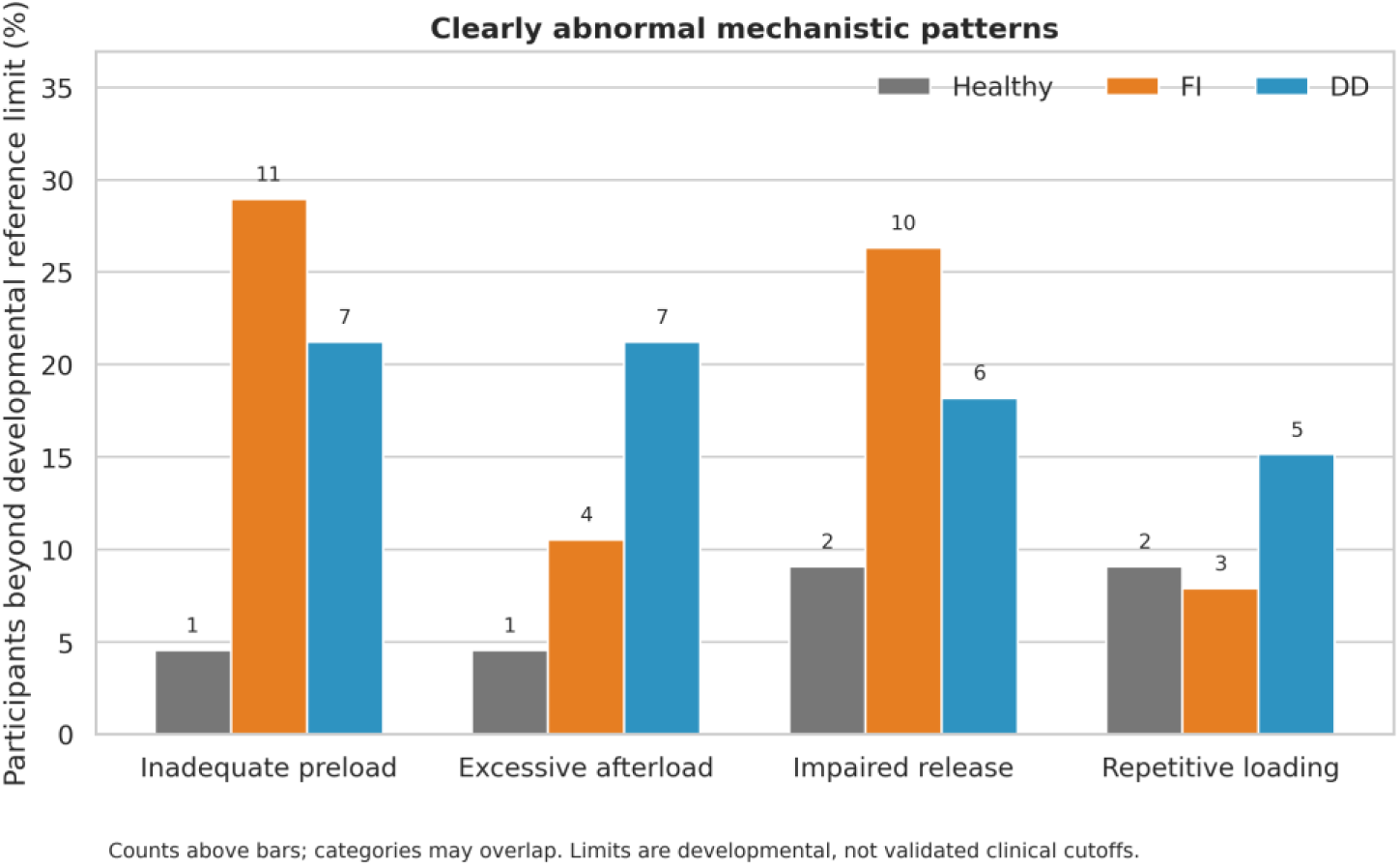
Proportions beyond developmental reference limits. Counts are shown above bars. Mechanisms may overlap, and overall group differences did not reach statistical significance. Defecatory disorder and fecal incontinence are abbreviated DD and FI.

### Construct validity and ARM outcomes

Within DD, worse constipation scores correlated with lower rear peak pressure (rho=-0.66), peak gradient (rho=-0.65), and pressure-space work rate (rho=-0.63); all Holm-adjusted P values were <0.001 and bootstrap intervals excluded zero **(Table 4**; **Figure 5).** These associations remained stable under winsorization and leave-one-out analysis.

**Figure 5.**
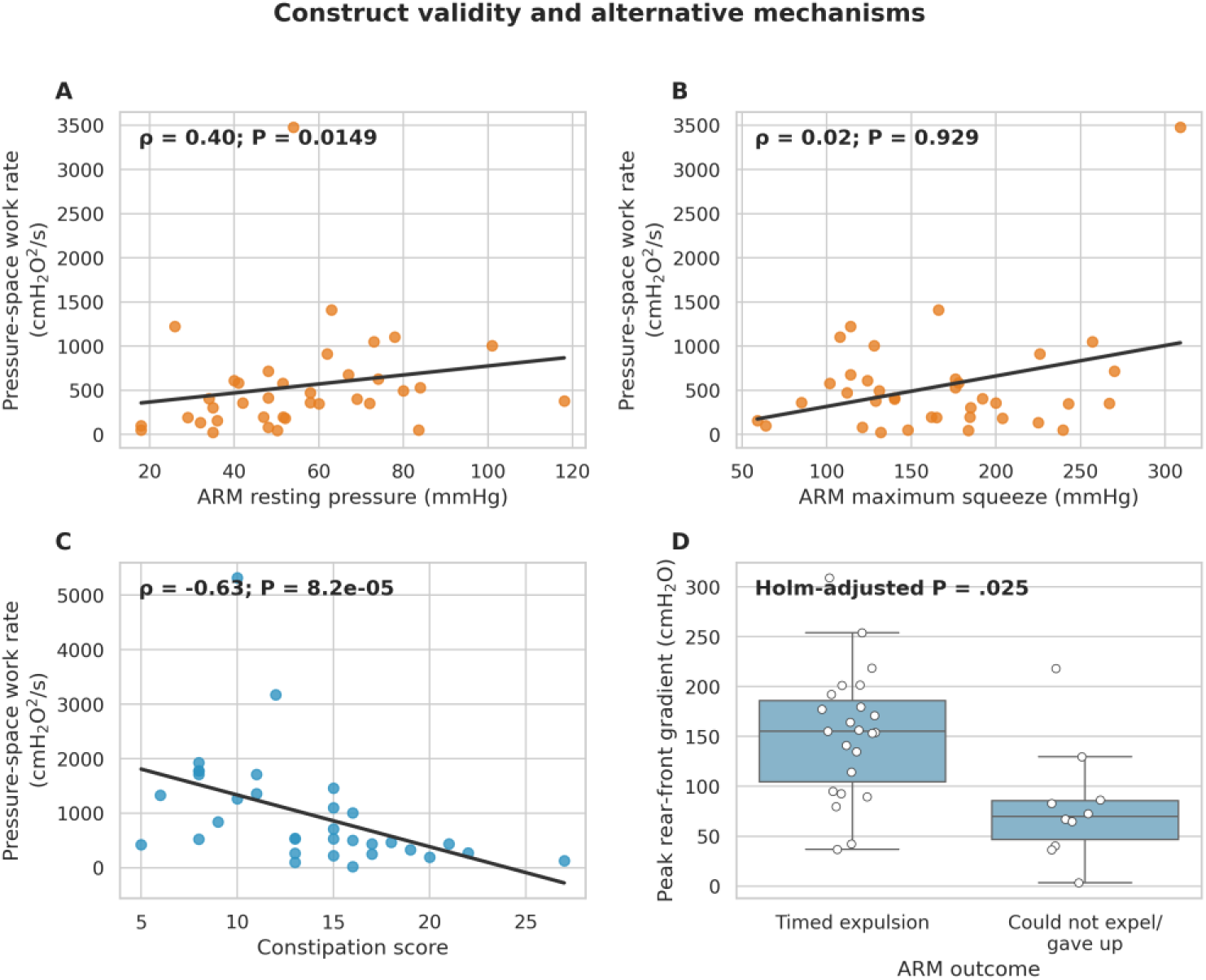
Construct validity and alternative mechanisms. A-B) FI pressure-space work rate versus ARM resting and maximum squeeze pressures. C) DD work rate versus constipation score. D) DD peak gradient according to ARM expulsion outcome. Correlations are Spearman coefficients; the P value in panel D is multiplicity adjusted.

**Table 4.**
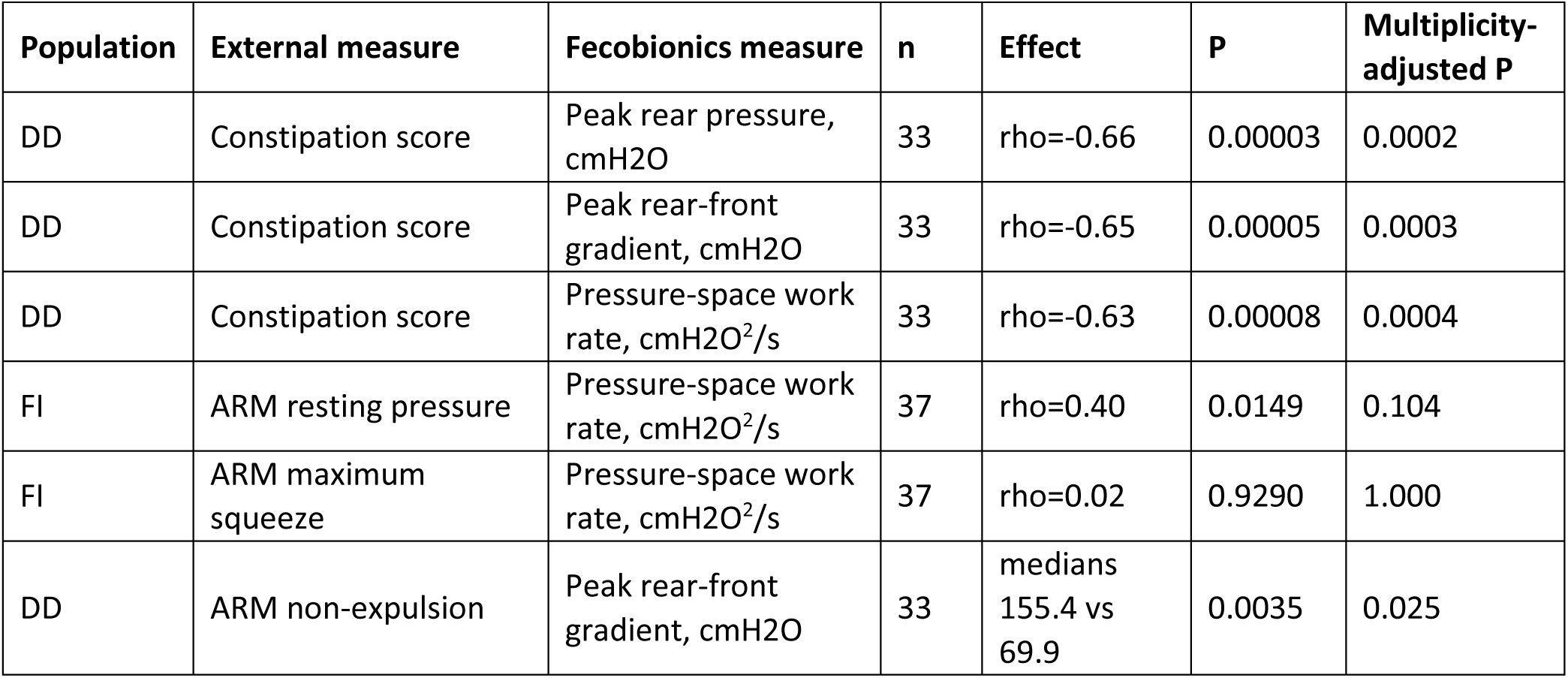
Construct-validity and competing-mechanism analyses.

| **Population** | **External measure** | **Fecobionics measure** | **n** | **Effect** | **P** | **Multiplicity-adjusted P** |
|---|---|---|---|---|---|---|
| DD | Constipation score | Peak rear pressure, cmH2O | 33 | rho=-0.66 | 0.00003 | 0.0002 |
| DD | Constipation score | Peak rear-front gradient, cmH2O | 33 | rho=-0.65 | 0.00005 | 0.0003 |
| DD | Constipation score | Pressure-space work rate, cmH2O<sup>2</sup>/s | 33 | rho=-0.63 | 0.00008 | 0.0004 |
| FI | ARM resting pressure | Pressure-space work rate, cmH2O<sup>2</sup>/s | 37 | rho=0.40 | 0.0149 | 0.104 |
| FI | ARM maximum squeeze | Pressure-space work rate, cmH2O<sup>2</sup>/s | 37 | rho=0.02 | 0.9290 | 1.000 |
| DD | ARM non-expulsion | Peak rear-front gradient, cmH2O | 33 | medians 155.4 vs 69.9 | 0.0035 | 0.025 |

ARM expulsion time was available for 23 DD patients; 10 additional patients were unable to evacuate within 120 seconds. Because missing timed values were informative, completer correlations were exploratory. Categorical non-evacuators had a lower median peak gradient than timed completers (69.9 versus 155.4 cmH2O; Holm-adjusted P=0.025).

Among 37 FI patients with ARM data, pressure-space work rate correlated nominally with resting pressure (rho=0.40, P=0.0149) but not squeeze pressure (rho=0.02, P=0.929). After correction across 10 FI-ARM tests, absolute phase work and peak gradient remained associated with resting pressure, whereas the primary work-rate association did not. Thus, reduced work output related to continence-system status but was not explained by maximal squeeze capacity.

In exploratory incremental analyses of the 33 DD examinations, the association between pressure-space work rate and constipation severity attenuated after adjustment for peak rear pressure and evacuation duration (partial Spearman rho = −0.17; bootstrap 95% confidence interval, −0.56 to 0.16; residual-permutation P = 0.378). Adding work rate did not improve leave-one-out prediction of constipation scores (root mean squared error, 4.79 versus 4.84 points). Analyses incorporating front-pressure amplitude or peak gradient, and alternative cohort definitions, likewise did not establish incremental association.

## Discussion

The principal finding was that clinically relevant abnormalities were expressed through different aspects of propulsion–outlet interaction in FI and DD. FI was characterized by reduced pressure-trajectory output, accompanied by lower rear pressure, peak rear–front gradient, and front-pressure release amplitude. DD showed substantial overlap with healthy participants at the group level, but greater constipation severity was associated with lower rear pressure, gradient, and pressure-space work rate. Patients unable to complete ARM-BET also had lower gradients. Together, these findings support the physiological relevance of achieved propulsion and its relationship to outlet loading. However, exploratory incremental analyses did not establish that pressure-space work rate contributed information about DD symptom severity beyond rear-pressure amplitude and evacuation duration.

### Defecatory disorder

In DD, the association between symptom severity and reduced rear pressure suggests that inadequate achieved propulsion contributes to dysfunction in at least a subset of patients. The lower gradient among ARM-BET non-evacuators provides complementary evidence that impaired evacuation can accompany an unfavorable balance between loading behind and ahead of the device. A low gradient can arise from insufficient rear pressure, persistent front pressure, or both; its components must therefore be examined before attributing failure to excessive outlet loading. These observations support a model in which clinically important DD includes inadequate pressurization as well as impaired outlet unloading.

This interpretation is consistent with simultaneous pressure and imaging studies showing contributions from inadequate rectal pressurization, insufficient anal opening, and disturbed abdominopelvic coordination.^1–6,28^ Our observations extend this physiological framework to pressures recorded along a stool-like device during attempted evacuation. They do not establish whether low rear pressure originates from reduced abdominal recruitment, altered rectal motor activity, pressure transmission, sensory factors, or the strategy adopted during the examination.

The relationship to the Rao classification^1–6,28^ requires explicit distinction. Inadequate propulsion and impaired anal relaxation are established mechanisms, and their recognition here is not itself novel. The additional question addressed by trajectory analysis is how loading and unloading interact across the complete attempt, including repeated efforts and terminal release. Similar peak pressures can coexist with different sequences of pressure change, while repeated large excursions can occur without successful evacuation. The potential contribution is therefore quantitative characterization of the evolution of dysfunction rather than replacement of established dyssynergia categories.

The incremental analyses qualify the interpretation of the symptom associations. Although pressure-space work rate correlated with constipation severity, this association attenuated after accounting for rear-pressure amplitude and evacuation duration, and adding work rate did not consistently improve internally validated prediction. These findings suggest substantial overlap between the information represented by the integral and simpler pressure measures. The small sample and wide confidence intervals limit precision, but the present results support interpreting work rate as a summary of pressure-trajectory behavior rather than an independently validated marker of dysfunction. Whether features specifically describing the timing of outlet unloading relative to propulsion provide additional information requires targeted evaluation.

Pressure-space work rate also requires physiological qualification. It measures pressure-plane excursion per unit time and is not mechanical power or a direct measure of evacuation efficiency. Its value depends on pressure amplitudes and the path traversed during repeated loading and unloading. A high value may accompany vigorous but unsuccessful cycling, whereas a low value may accompany limited pressurization or a relatively low-load evacuation. It should therefore be interpreted jointly with gradient, release behavior, and evacuation outcome.

### Fecal incontinence

In FI, reduced pressure output during intentional evacuation raises the possibility that impaired continence and evacuation behavior are functionally linked. One explanation is altered sensorimotor function or pressure transmission that reduces the pressure generated around the device. An alternative is reduced recruitment of abdominal effort by patients who associate increased pressure with leakage. These physiological and behavioral explanations can coexist: reduced continence reserve could influence the motor strategy selected during evacuation. The present study demonstrates reduced achieved output under the examination conditions; it does not establish reduced maximal propulsive capacity or anticipatory suppression of effort. The present protocol did not measure maximal abdominal force, motivation, perceived leakage threat, or fear during attempted evacuation. Reduced output may reflect impaired propulsion, altered pressure transmission, reduced rectal loading, or low continence-system reserve. It may also be adaptive: patients who have learned that increased abdominal pressure precipitates leakage may consciously or subconsciously restrict straining during the test. Psychological and avoidance factors are recognized components of the burden and management of incontinence, but they were not measured here.^8–10,14,15,29^ This needs further study.

The ARM associations provide limited support for this interpretation. Associations of absolute pressure-space work and peak gradient with resting pressure survived multiplicity adjustment, whereas the work-rate association did not. The absence of an association with maximal squeeze pressure does not exclude muscular dysfunction because anal squeeze and evacuation involve different motor tasks and muscle contributions. Consequently, these findings cannot distinguish shared anorectal impairment from adaptation to reduced continence reserve. Habitual and standardized maximal efforts, combined with abdominal recruitment measurements and assessment of expected leakage, would help discriminate these explanations.

Reduced front-pressure release amplitude in FI should likewise be interpreted in relation to the pressure available to decline. A smaller absolute fall can follow a lower initial pressure without indicating defective relaxation. In addition, front pressure changes as the device moves and its contact with surrounding tissues changes. The observed release measure therefore describes unloading at the device front and cannot be assigned exclusively to sphincter relaxation. Analyses conditional on preceding front pressure, together with temporal alignment to passage, would help distinguish reduced loading from impaired unloading.

The developmental phenotype categories provide a framework for formulating intervention hypotheses. Low rear-pressure patterns suggest testing whether improved recruitment increases achieved propulsion; persistent front loading suggests testing strategies directed at outlet relaxation and coordination. Repetitive loading suggests examining whether effort is appropriately timed to unloading. These are predictions for prospective studies rather than established treatment indications. Demonstrating that a targeted intervention changes the predicted trajectory feature and improves evacuation would provide stronger mechanistic evidence than phenotype assignment alone.

### Reference limits and clinical translation

The fifth/95th-percentile limits were intentionally specific and identified only clearly abnormal trajectories. They are unsuitable for screening. The normative sample of 20 produces wide percentile uncertainty, and some apparently HP may still have occult dysfunction. Reference limits should be stratified or modeled continuously in larger cohorts and validated on a separate device and population before diagnostic use.^1–4^

### Strengths and limitations

Strengths include simultaneous axial pressure recording, analysis of complete trajectories, preservation of sustained and repetitive events, channel-specific rather than subject-wide exclusion, objective recovery of additional intervals, and consistent findings across cohort tiers and robust sensitivity analyses. The analysis also separated timed ARM completion from non-expulsion rather than treating missing time as random.

The primary limitations of this study include its single-center nature, a relatively homogeneous study population consisting mostly of Chinese individuals, retrospective design, a small normative sample, incomplete clinical measurements, uncertain correspondence between device pressure and specific muscle groups, and lack of direct abdominal force, fear, or behavioral measures. The recovered intervals require final blinded confirmation. Phenotype rules were developed and evaluated in the same cohort, prevalence differences were not significant, and no external validation was available. Multiple trajectory variables are correlated, and exploratory results remain vulnerable to optimism. The front-channel failure reduced the paired-pressure cohort by one DD examination.

### Conclusions

Fecobionics preload-afterload metrics identified reduced pressure-work output in FI and heterogeneous failure patterns in DD. Reduced output in FI should not be equated with reduced physiological capacity; it may combine low continence-system reserve with anticipatory suppression of abdominal effort. DD severity was associated with lower preload, gradient, and work rate, while ARM-BET non-evacuators had particularly low-pressure gradients. Developmental mechanistic phenotypes require prospective adjudication and external validation.

## Data Availability

All data produced in the present study are available upon reasonable request to the authors

## Acknowledgements

Research reported in this publication was supported by the National Institutes of Health (USA) under Award Number R01DK125657, R01DK131488, and R01DK134689. The Chinese University of Hong Kong (strategic recruitment funding), RCG grant #14106717, and the National Natural Science Foundation of China grant # 11902057. Dr. K Futaba, Dr. SC Chen, Mr. WW Leung, and Ms. C Wong are kindly thanked for obtaining the data.

## Data availability statement

Access to data can be granted upon reasonable request, which should be directed to the corresponding author.

## Author contributions

HG designed the study and advised the experimental work during the study period. HG participated in most experiments. Data analysis was conducted by DS, JY, LK and HG and interpreted by all authors. All authors had access to the data, revised the manuscript and approved the final version for submission. HG is guarantor of the article.

## Supplementary Material

### Supplementary Methods

#### Notation and interpretation

E denotes the analyzed evacuation interval and T its duration. Pfront and Prear are baseline-corrected front and rear pressures. Pressure-space work has units of pressure squared and is a phase-plane descriptor, not physical work or energy.

#### Absolute pressure-space work

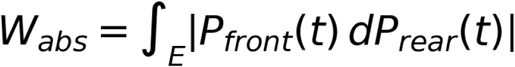

The absolute line integral counts repeated traversal of pressure space without cancellation and therefore represents total cyclic pressure interaction.

#### Pressure-space work rate

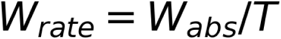

The prespecified primary endpoint divides absolute pressure-space work by analyzed interval duration.

#### Signed work and hysteresis ratio

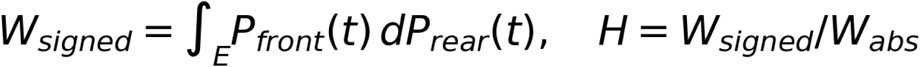

Signed work retains trajectory direction. H normalizes signed work to total absolute traversal and ranges theoretically from -1 to 1.

#### Active afterload fraction

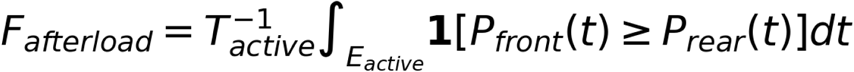

Active effort was defined by smoothed rear pressure exceeding the larger of 10 cmH2O or 20% of its peak. The indicator is one when front pressure equals or exceeds rear pressure.

#### Pressure-cycling index

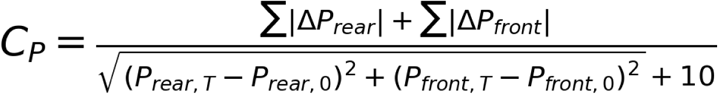

Total front and rear pressure variation was divided by net pressure-plane displacement plus a 10-cmH2O stabilizer approximating a conservative physiological noise scale.

#### Landmarks and smoothing

A centered 0.35-second rolling median identified rear and front peaks and contraction landmarks. Reported pressure integrals used the processed pressure trajectory after baseline correction and the limited isolated-spike rule. Sustained elevations were not removed because of amplitude. Rear contractions required prominence of at least 10 cmH2O or three baseline-noise units and separation of at least one second.

#### Interval hierarchy

Tier 1 contained conventional investigator-marked intervals. Tier 2 added investigator-created analysis blocks with explicit time columns, row alignment, or unique sequence matching. Tier 3 added conservatively reviewed terminal intervals with coordinated movement and terminal pressure unloading. The expanded Tier 2 cohort was primary; Tiers 1 and 3 were sensitivity cohorts.

#### Reference limits and uncertainty

The normative subset contained 20 unqualified healthy participants with symptom scores inside the prespecified normal ranges. Empirical fifth and 95th percentiles were accompanied by percentile bootstrap intervals from 4,000 resamples. A regularized four-axis covariance model estimated an exploratory multivariable reference distance using rear peak, active-afterload fraction, release amplitude, and cycling index. Its internal 95th-percentile distance was not used as a diagnostic cutoff.

#### Robust and missing data analyses

Rank-test sensitivity used first/99th-percentile winsorization without deleting participants. Correlations were checked using five-percent winsorization, bootstrap intervals, and leave-one-out ranges. ARM expulsion time was missing informatively when participants could not expel or gave up; timed-completer correlations and categorical non-expulsion comparisons were therefore reported separately.

#### Blinded technical review

Boundary and extreme cases were selected by numerical distance from each developmental limit without use of diagnosis. Review images showed time-pressure traces and phase trajectories under anonymous case identifiers. The technical review identified isolated spikes but did not perform clinical or physiological adjudication. The diagnosis key remained separate from the scoring sheet.

## Supplementary Tables

**Table S1.** Complete univariate trajectory analysis (selected statistical columns).

| **Measure** | **Healthy n** | **Healthy median** | **Healthy Q1** | **Healthy Q3** | **FI median** | **DD median** | **Overall P** | **FI AUC** | **DD AUC** |
|---|---|---|---|---|---|---|---|---|---|
| peak_gradient | 22 | 140.393 | 119.399 | 186.358 | 95.754 | 134.657 | 0.004 | 0.76 | 0.583 |
| phase_absolute_work | 22 | 19316.4 | 12189.317 | 39591.807 | 11586.326 | 23227.713 | 0.005 | 0.694 | 0.51 |
| phase_work_rate | 22 | 803.492 | 523.329 | 1135.791 | 389.348 | 528.815 | 0.006 | 0.732 | 0.545 |
| front_release_amplitude | 22 | 98.354 | 61.554 | 130.527 | 58.635 | 85.353 | 0.007 | 0.708 | 0.578 |
| front_peak | 22 | 54.709 | 20.522 | 90.719 | 29.675 | 57.106 | 0.03 | 0.648 | 0.51 |
| rear_peak | 22 | 112.784 | 93.867 | 146.321 | 67.507 | 107.583 | 0.032 | 0.715 | 0.596 |
| phase_path_length | 22 | 947.734 | 754.611 | 3170.67 | 877.921 | 1365.316 | 0.062 | 0.614 | 0.556 |
| terminal_front_residual | 22 | -35.436 | -47.533 | -18.279 | -23.276 | -35.181 | 0.08 | 0.669 | 0.58 |
| mean_active_gradient | 22 | 70.896 | 42.991 | 87.503 | 46.52 | 42.974 | 0.155 | 0.654 | 0.618 |
| minimum_gradient | 22 | -19.426 | -47.571 | -11.179 | -30.669 | -36.099 | 0.155 | 0.581 | 0.647 |
| phase_signed_work | 22 | 4110.628 | -203.437 | 8656.312 | 3165.735 | 5231.66 | 0.22 | 0.533 | 0.576 |
| active_afterload_fraction | 22 | 0.005 | 0.0 | 0.129 | 0.0 | 0.035 | 0.24 | 0.547 | 0.581 |
| pressure_cycling_index | 22 | 18.268 | 12.667 | 46.178 | 15.914 | 24.588 | 0.247 | 0.602 | 0.539 |
| release_slope_front_on_rear | 22 | 0.287 | 0.001 | 0.407 | 0.147 | 0.218 | 0.55 | 0.542 | 0.517 |
| phase_hysteresis_ratio | 22 | 0.235 | 0.021 | 0.378 | 0.228 | 0.262 | 0.622 | 0.51 | 0.561 |
| phase_path_per_peak_sum | 22 | 8.472 | 4.991 | 12.043 | 9.221 | 9.066 | 0.666 | 0.559 | 0.566 |
| front_peak_delay_s | 22 | -6.464 | -11.081 | -1.57 | -4.823 | -3.199 | 0.721 | 0.577 | 0.53 |
| contraction_count | 22 | 3.5 | 2.0 | 7.0 | 2.5 | 3.0 | 0.749 | 0.554 | 0.501 |
| phase_work_per_peak_product | 22 | 4.706 | 2.306 | 11.019 | 4.103 | 6.116 | 0.787 | 0.507 | 0.54 |
| loading_slope_front_on_rear | 22 | 0.054 | -0.254 | 0.478 | 0.031 | 0.03 | 0.974 | 0.52 | 0.511 |

**Table S2.** Developmental healthy reference limits with bootstrap intervals.

| **Measure** | **Normative n** | **P05** | **P05 CI low** | **P05 CI high** | **P95** | **P95 CI low** | **P95 CI high** | **Median** | **Q1** | **Q3** |
|---|---|---|---|---|---|---|---|---|---|---|
| rear_peak | 20 | 43.747 | 32.122 | 72.336 | 181.999 | 148.679 | 198.952 | 112.784 | 87.494 | 144.931 |
| front_peak | 20 | 3.174 | -15.959 | 16.418 | 140.572 | 101.669 | 226.791 | 54.709 | 18.789 | 93.753 |
| peak_gradient | 20 | 84.853 | 42.421 | 118.711 | 216.757 | 188.184 | 237.294 | 140.393 | 121.617 | 183.605 |
| minimum_gradient | 20 | -68.318 | -85.659 | -49.483 | 4.719 | -8.77 | 13.002 | -19.426 | -48.03 | -10.376 |
| active_afterload_fraction | 20 | 0.0 | 0.0 | 0.0 | 0.304 | 0.142 | 0.34 | 0.005 | 0.0 | 0.157 |
| mean_active_gradient | 20 | 12.344 | 11.457 | 40.749 | 108.371 | 89.568 | 121.027 | 67.857 | 42.014 | 89.494 |
| phase_signed_work | 20 | -25567.725 | -84855.512 | -655.993 | 24660.232 | 8834.963 | 55991.745 | 3999.789 | -430.767 | 8421.611 |
| phase_absolute_work | 20 | 1925.62 | 1097.061 | 11663.674 | 324534.621 | 87885.451 | 361582.062 | 17806.747 | 11836.591 | 51931.735 |
| phase_hysteresis_ratio | 20 | -0.236 | -0.255 | -0.021 | 0.744 | 0.406 | 0.855 | 0.235 | -0.011 | 0.321 |
| phase_path_length | 20 | 566.011 | 530.213 | 688.034 | 15714.317 | 3260.235 | 18775.591 | 910.237 | 734.563 | 3014.81 |
| phase_work_rate | 20 | 118.311 | 54.768 | 518.593 | 3927.625 | 1214.102 | 4188.212 | 833.388 | 524.77 | 1173.282 |
| phase_work_per_peak_product | 20 | 0.987 | 0.738 | 1.956 | 22.533 | 11.269 | 25.076 | 4.706 | 2.011 | 10.773 |
| phase_path_per_peak_sum | 20 | 3.434 | 3.374 | 4.298 | 40.249 | 12.423 | 80.507 | 8.037 | 4.472 | 11.881 |
| pressure_cycling_index | 20 | 8.677 | 6.841 | 11.68 | 177.072 | 47.766 | 392.481 | 18.193 | 11.936 | 43.485 |
| loading_slope_front_on_rear | 20 | -0.928 | -1.458 | -0.36 | 0.776 | 0.593 | 0.785 | 0.044 | -0.282 | 0.512 |
| release_slope_front_on_rear | 20 | -0.091 | -0.093 | -0.06 | 0.691 | 0.412 | 0.883 | 0.287 | -0.019 | 0.41 |
| front_peak_delay_s | 20 | -14.22 | -28.132 | -11.013 | 1.321 | -0.628 | 13.902 | -5.969 | -10.271 | -1.147 |
| contraction_count | 20 | 0.95 | 0.0 | 1.95 | 18.65 | 7.1 | 50.0 | 2.5 | 2.0 | 6.25 |
| terminal_front_residual | 20 | -78.439 | -86.576 | -50.276 | 0.056 | -18.177 | 1.122 | -35.436 | -49.292 | -18.33 |
| front_release_amplitude | 20 | 14.74 | 4.181 | 63.227 | 179.557 | 138.094 | 245.045 | 98.354 | 64.741 | 137.538 |

**Table S3.**
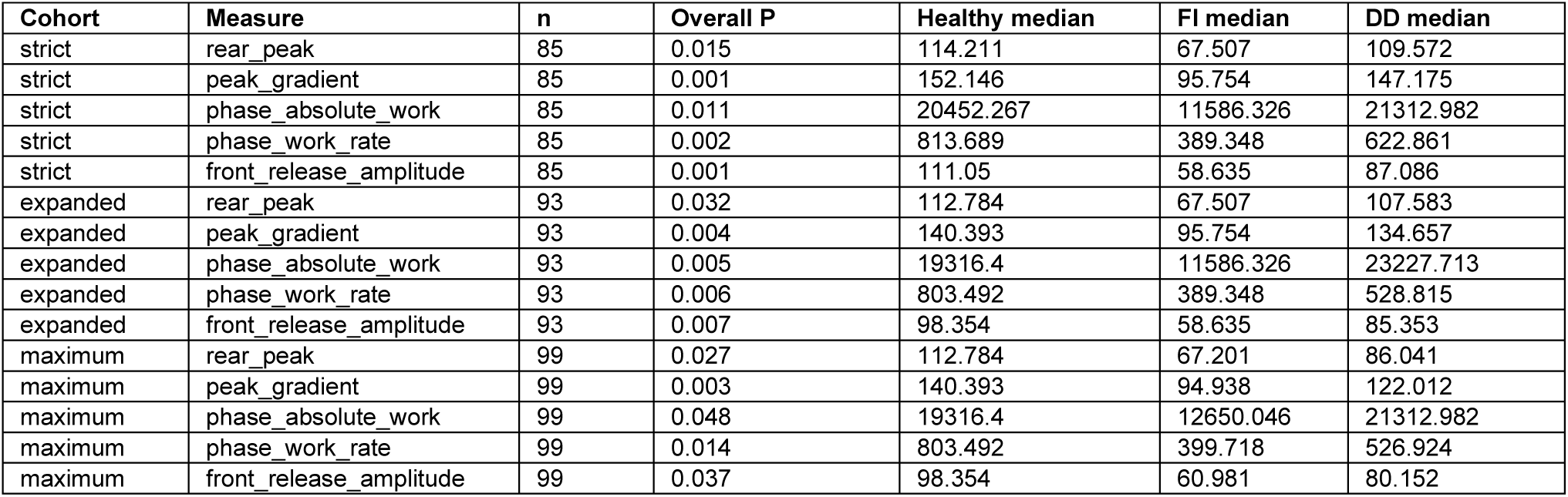
Cohort-tier sensitivity for primary and key secondary endpoints.

**Table S4.**
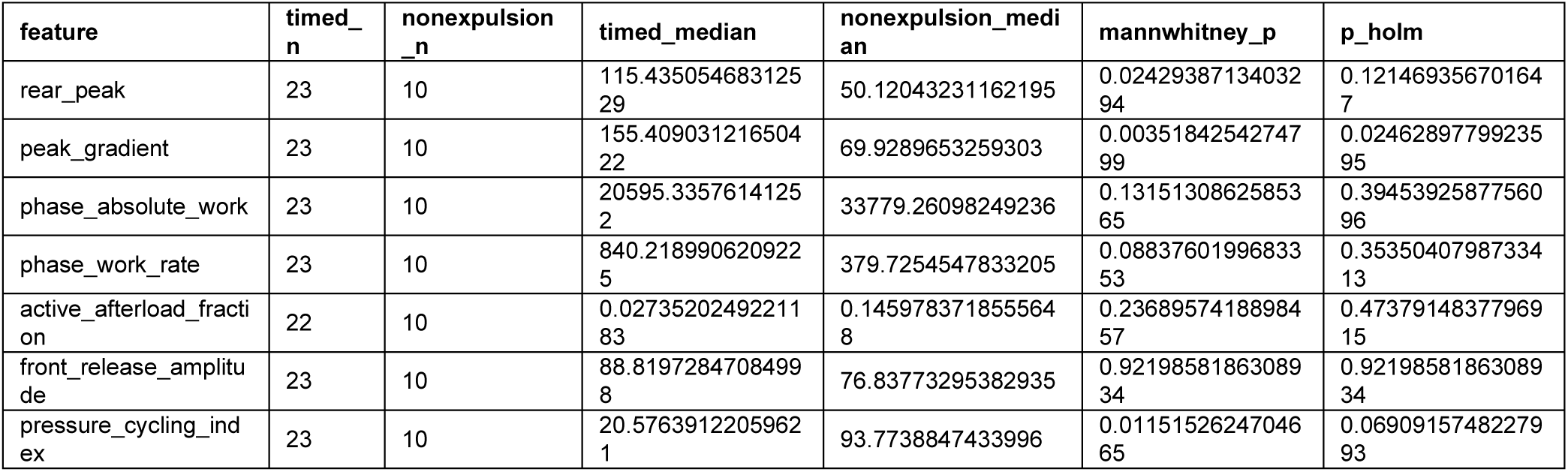
Preload-afterload measures according to ARM timed expulsion versus non-expulsion.

| feature | timed_n | nonexpulsion_n | timed_median | nonexpulsion_median | mannwhitney_p | p_holm |
|---|---|---|---|---|---|---|
| rear_peak | 23 | 10 | 115.43505468312529 | 50.12043231162195 | 0.0242938713403294 | 0.121469356701647 |
| peak_gradient | 23 | 10 | 155.40903121650422 | 69.9289653259303 | 0.0035184254274799 | 0.0246289779923595 |
| phase_absolute_work | 23 | 10 | 20595.3357614125​2 | 33779.26098249236 | 0.1315130862585365 | 0.3945392587756096 |
| phase_work_rate | 23 | 10 | 840.2189906209225 | 379.7254547833205 | 0.0883760199683353 | 0.3535040798733413 |
| active_afterload_fraction | 22 | 10 | 0.0273520249221183 | 0.1459783718555648 | 0.2368957418898457 | 0.4737914837796915 |
| front_release_amplitude | 23 | 10 | 88.81972847084998 | 76.83773295382935 | 0.9219858186308934 | 0.9219858186308934 |
| pressure_cycling_index | 23 | 10 | 20.5763912205962​1 | 93.7738847433996 | 0.0115152624704665 | 0.0690915748227993 |

**Table S5.**
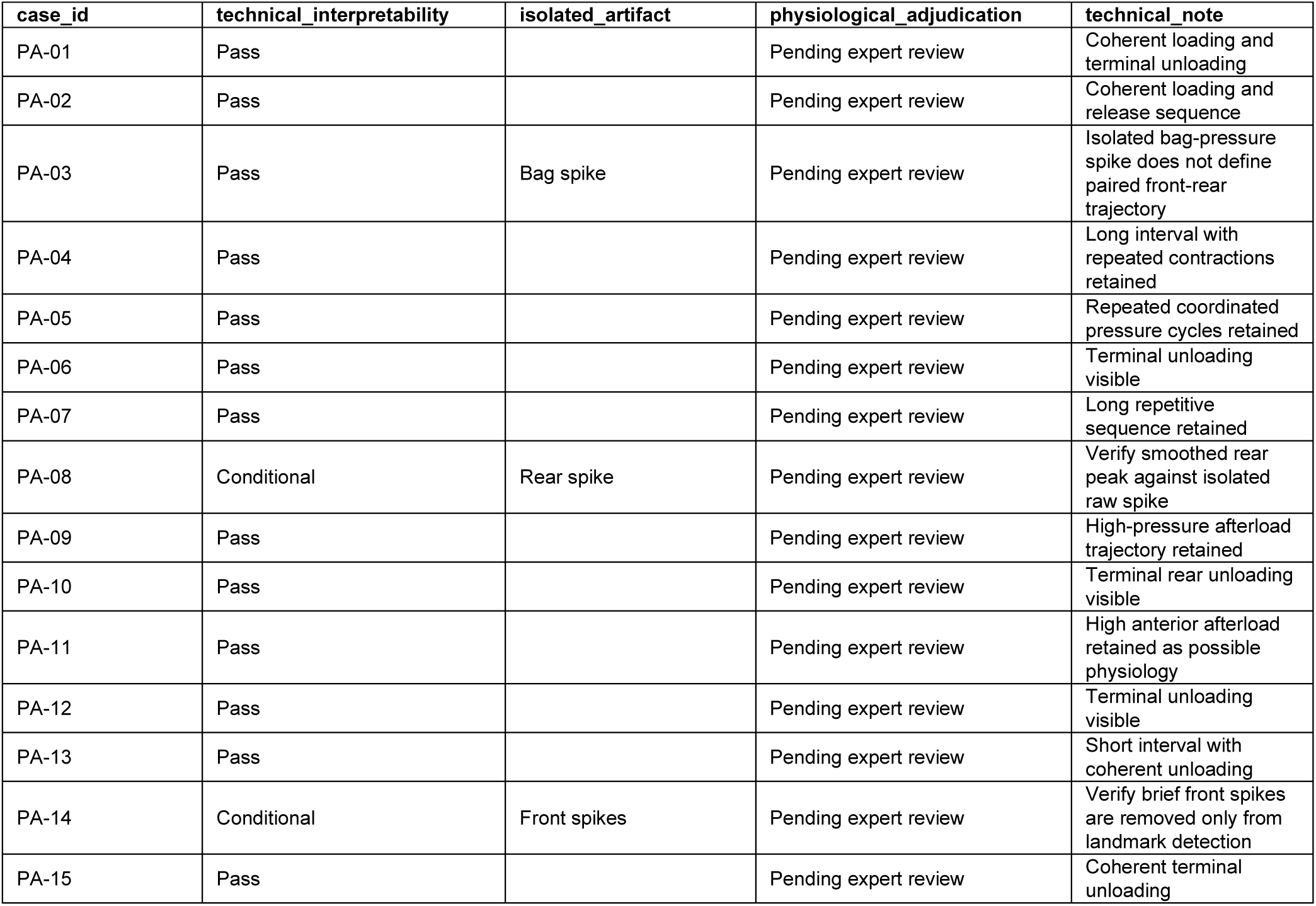
Diagnosis-blinded technical boundary review.

